# Construction of a demand and capacity model for intensive care and hospital ward beds, and mortality from COVID-19

**DOI:** 10.1101/2021.01.06.21249341

**Authors:** Stuart McDonald, Chris Martin, Steve Bale, Michiel Luteijn, Rahul Sarkar

## Abstract

**Background:** This paper describes the construction of a model used to estimate the number of excess deaths that could be expected as a direct consequence of a lack of hospital bed and intensive care unit (ICU) capacity.

**Methods:** A series of compartmental models was used to estimate the number of deaths under different combinations of care required (ICU or ward), and care received (ICU, ward or no care) in England up to the end of April 2021. Model parameters were sourced from publicly available government information, organisations collating COVID-19 data and calculations using existing parameters. A compartmental sub-model was used to estimate the mortality scalars that represent the increase in mortality that would be expected from a lack of provision of an ICU or general ward bed when one is required. Three illustrative scenarios for admissions numbers, ‘Optimistic’, ‘Middling’ and ‘Pessimistic’, are described showing how the model can be used to estimate mortality rates under different scenarios of capacity.

**Results:** The key output of our collaboration was the model itself rather than the results of any of the scenarios. The model allows a user to understand the excess mortality impact arising as a direct consequence of capacity being breached under various scenarios or forecasts of hospital admissions. The scenarios described in this paper are illustrative and are not forecasts.

There were no excess deaths from a lack of capacity in any of the ‘Optimistic’ scenario applications in sensitivity analysis.

Several of the ‘Middling’ scenario applications under sensitivity testing resulted in excess deaths directly attributable to a lack of capacity. Most excess deaths arose when we modelled a 20% reduction compared to best estimate ICU capacity. This led to 597 deaths (0.7% increase).

All the ‘Pessimistic’ scenario applications under sensitivity analysis had excess deaths. These ranged from 49,219 (19.4% increase) when we modelled a 20% increase in ward bed availability over the best-estimate, to 103,845 (40.9% increase) when we modelled a 20% shortfall in ward bed availability below the best-estimate. The emergence of a new, more transmissible variant (VOC 202012/01) increases the likelihood of real world outcomes at, or beyond, those modelled in our ‘Pessimistic’ scenario.

The results can be explained by considering how capacity evolves in each of the scenarios. In the Middling scenario, whilst ICU capacity may be approached and even possibly breached, there remains sufficient ward capacity to take lives who need either ward or ICU support, keeping excess deaths relatively low. However, the Pessimistic scenario sees ward capacity breached, and in many scenarios for a period of several weeks, resulting in much higher mortality in those lives who require care but do not receive it.

**Conclusions:** No excess deaths from breaching capacity would be expected under the unadjusted ‘Optimistic’ assumptions of demand. The ‘Middling’ scenario could result in some excess deaths from breaching capacity, though these would be small (0.7% increase) relative to the total number of deaths in that scenario. The ‘Pessimistic’ scenario would certainly result in significant excess deaths from breaching capacity. Our sensitivity analysis indicated a range between 49,219 (19.4% increase) and 103,845 (40.9% increase) excess deaths.

Without the new variant, exceeding capacity for hospital and ICU beds did not appear to be the most likely outcome but given the new variant it now appears more plausible and, if so, would result in a substantial increase in the number of deaths from COVID-19.

## Background

The number of deaths from COVID-19 in the UK was 74,125 deaths and the number of known cases was 2,542,069 as of the 2^nd^ January 2021 representing a case fatality rate (CFR) of 2.9%. The Office for National Statistics (ONS) Coronavirus (COVID-19) Infection Survey estimated that 8.7% of people in England still had antibodies at detectable levels based upon serological testing.(1) Assuming that, as of the 2^nd^ January 2021, around 10% to 15% of the population of England has been infected (taking into account those with antibodies no longer at detectable levels, the lag time and the small proportion of false negative serology), then this would suggest that between 6.7 million and 10 million people have already been infected representing an infection fatality rate (IFR) of 0.7% to 1.1%.

IFR estimates are typically made in the context of adequate capacity of health care services including hospital beds and intensive care unit (ICU) beds. Should the demand for ICU beds exceed the supply, then the IFR would be expected to rise. In this paper we describe a demand and capacity model designed to estimate the number of deaths that would directly arise from a lack of ICU and ward beds.

In November 2020 we began development of a model to estimate what increases in deaths could be expected as a direct consequence of a lack of hospital bed capacity, and in particular ICU beds. Due to availability of data, the model was restricted to England.

Estimating the ICU capacity in England is not straightforward. There were estimated to be 4,114 ICU beds in England pre-pandemic.(2) However, there are plans in place to allow surges in ICU capacity when pandemics occur which entails repurposing other hospital resources including anaesthetic rooms, operating theatres and parts of accident and emergency departments. The aim was to be able to increase ICU bed capacity across the country by 100% in the event of a pandemic.(3) It is likely that the increase in capacity will vary by institution and one case study managed to increase capacity by 236% during the first wave.(4) In the event of a pandemic, the NHS is supposed to respond through a whole system approach, which has been outlined in the UK CRITCON scoring system. The basis for the system is that the same non-pandemic ethical standards are applied to treat patients and allocate resources, unless in the extreme scenario (CRITCON 4). The system was formulated in 2009 for H1N1 and has been updated in 2020 in the context of the COVID-19 pandemic. The levels described above extend from a normal capacity at CRITCON 0 to CRITCON 4 when the system is overwhelmed. A “mutual aid” system exists to facilitate inter-hospital or regional transfer of patients when the local intensive care capacity is breached in order to support the principle that no patient should be deprived of the appropriate care if there is systemwide capacity available. Eighteen intensive care networks in the England and Northern Ireland manage the capacity and allocation to intensive care in the event of increasing demand.

At the start of the pandemic a series of field hospitals were constructed at eight sites across England. The capacity is somewhat elastic according to circumstances, but it is estimated that this would potentially provide an additional 8,000 general ward beds and 500 ICU beds though there may be staffing constraints that also limit this number.(2)

In this paper we describe a model for estimating the number of additional deaths that occur from a lack of capacity under different scenarios. These are illustrative scenarios and not forecasts. It is important to emphasise that parameterization of the model can be modified according to available information and updated over time.

## Methods

### Model description

The model can be found at this location on GitHub: Crystallize/COVID19_ExceedingCapacityModel (github.com).

The model is a series of static compartmental models that estimates the number of deaths under three conditions: availability of both general ward and ICU care; availability of general ward care but no ICU care; and no availability of either general ward or ICU care.

The model was developed as an Excel workbook. This was chosen as it allows rapid iteration and development with transparency for other developers and reviewers as it is a widely available platform.

### General outline

The general outline of the model is shown in Figure 1. It operates using a scenario of COVID hospital admission demand in weekly time steps.

**Figure 1.**
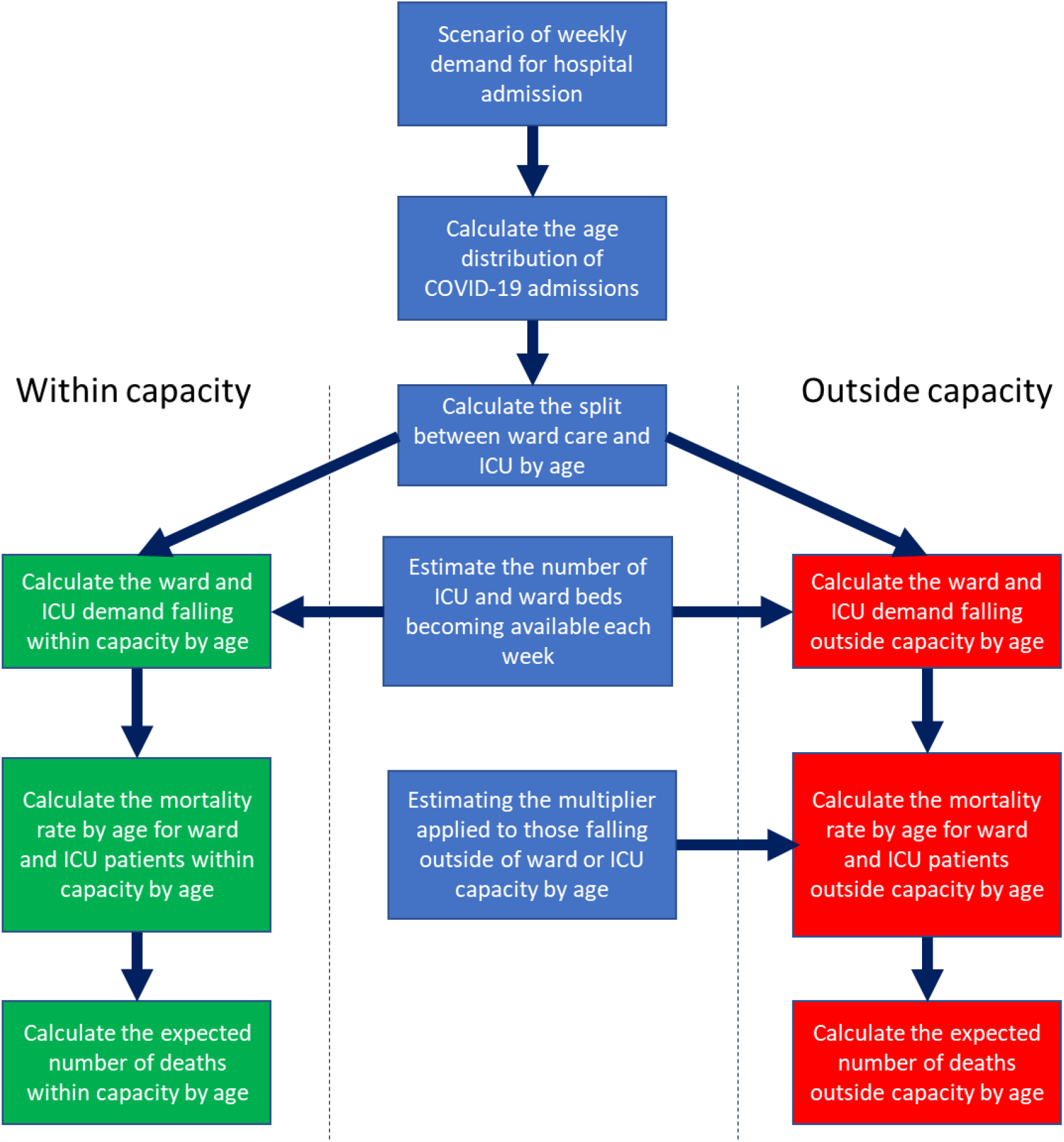
Outline of the structure of the model.

First, we modelled the expected age distribution of admissions. This was important as hospital and intensive care admission rates and mortality varies substantially by age. Subsequently, the weekly demand for intensive care and ward beds was modelled by age. The weekly availability of beds was calculated using estimates of the number of free beds usually available in ICUs in England plus any surge capacity and additional capacity freed by the cancellation of routine surgery, field hospitals and the use of private facilities adjusted for the average duration of stay on ICU and in the ward in general.

We then compared the weekly demand for ward and ICU beds with the maximum number of beds available. This allowed for the number of weekly admissions that fall within or outside of capacity.

Next, we calculated an estimate of the multiplier of the mortality rates when an ICU bed is not available to someone who needs it, and similarly for general ward care. This was done by compartmentalizing ward and ICU patients to categories of care for which estimates were made and then aggregated up to the ward or ICU level again. The assumption is that the ICU mortality rate is multiplied by 1.99 if there is no ICU bed available, but there is a ward bed, and by 9.02 if there is neither a ward not ICU bed available. For general ward patients, the multiplier is 3.69 if there are no ward beds available. More detail on the derivation of this is provided in the section on the “Excess Mortality Model” below. Sensitivity testing indicated that results were not especially sensitive to small changes in these mortality multipliers (+/-20%).

Once mortality rates for ward and ICU patients both within and outside of capacity were calibrated, the number of deaths occurring ‘within capacity’ and the number that occur directly as a result of being out of capacity could be calculated. The number of ‘out of capacity’ deaths does not include the number of deaths that would have been expected to occur if normal care had been received, and so represent ‘excess’ deaths occurring purely because the capacity limit was breached.

### Description of each step in the modelling

We will now describe the purpose and workings of the model in more detail.

### Principle parameters

The pre-pandemic (i.e. before March 2020) spare bed availability in the general wards is estimated at 9,769 and the ICU spare bed capacity as 817 beds from a study of hospital capacity in the COVID-19 pandemic by the Medical Research Council, Public Health England, The National Institute of Health Research and Imperial College.(2) The same study estimated an additional 1,810 ICU beds and 52,498 beds could be acquired using field hospitals, the cancellation of routine care and the use of private hospital facilities. In addition, there are pandemic response plans in place to increase ICU capacity with a surge in demand. This surge capacity would be intended to increase ICU beds by 100% by using anaesthetic rooms, operating theatres and other hospital resources.(3) Altogether this would increase spare ICU capacity to 3,444 beds and spare ward capacity to 62,267 beds. These 62,267 ward beds can provide for 87,174 patients weekly. These figures are based on an estimated length of stay on ICU of 7 days and length of stay on the general ward of 5 days.(5,6)

### Estimating Admissions by Age

For this initial step, the age distribution of admissions was required in 5-year age bands up to the age of 80 years with one category for those 80-years and over. If this data is available directly, it can be applied in the model. In ICUs, the age distribution is lower with an average age of 62 years and with only a small proportion of over 80s being fit enough for invasive ventilation.(5) However, we estimated the distribution of admissions for each age-band for each calendar week by fitting an exponential curve to age-binned data on admission rates for adults, then interpolating admission rates for each year of age. Using population estimates from 2019 for each year of age, estimates of the numbers of admissions were made. These were then recalibrated so that the exact number of admissions in each age group matched the observed numbers in each corresponding bin.

### New patients in hospital

In this step, we applied the distribution of admissions by age to the weekly admission demand from the scenarios to calculate the percentage of admissions accounted for by each year of age.

### Splitting Admissions into ICU and Ward

Next, we calculated the proportion of admissions that require intensive care and ward care by age for each 5-year age band, with one age bin for those aged 80 or over. This is done using data on the age breakdown of admissions and total admission numbers to ICU from the ICNARC report of the 18^th^ December 2020, and the total number of COVID-19 hospital admissions up to the 18^th^ December 2020 from the Gov.UK COVID-19 data dashboard.(7,8)

### Ward Demand

In this step, we separated the weekly demand for general ward care by age into two tables; one for those cared for within the expected hospital capacity, and a second for those who fail to receive any hospital care when required because of no capacity.

### ICU Demand

In this step we separated the weekly demand for ICU care by age into three tables; one for those cared for within the expected ICU capacity, a second for those who receive only ward care when ICU care is required because of no capacity, and a third for those who fail to receive any hospital care when required.

### Excess Mortality Model

In this step of the model, we estimated the multiplier of mortality risk for ward and ICU care COVID-19 patients when there is 1) no ICU capacity and 2) when there is neither ward nor ICU capacity. The output from this sub-model is only dependent on the ‘hospitalised’ compartment and the modelling is not age dependent. The proportion of patients requiring intensive care was taken from the results of the modelling step “Splitting Admissions into ICU and Ward” (8%). Those receiving general ward care or ICU care were segmented into the categories shown in Table 1. The distribution of ward care patients across the care categories was populated using observation data from the Nottingham Universities Hospitals Trust.(9) The proportion of ICU patients only requiring supportive care or high flow oxygen was also take from the Nottingham data. The rest of the ICU categories were populated using data from Southampton.(10)

**Table 1.** Hospital care compartments, proportion occupancy and the mortality rates under three capacity scenarios

| Proportion | Care segment | Proportion in each group | Mortality: in capacity | Mortality: no ICU | Mortality: no ward or ICU |
| --- | --- | --- | --- | --- | --- |
| 92% | General ward care | 0.6 | 0.01 | 0.01 | 0.03 |
|  | Ward care: O<sub>2</sub>>35% | 0.17 | 0.01 | 0.01 | 0.5 |
|  | Ward care: O<sub>2</sub>>35% (Ceiling) | 0.23 | 0.4 | 0.4 | 0.95 |
| 8% | ICU: Supportive/HFO | 0.48 | 0.01 | 0.02 | 0.75 |
|  | ICU: NIV/CPAP | 0.16 | 0.01 | 0.02 | 0.95 |
|  | ICU: NIV/CPAP (Ceiling) | 0.12 | 0.83 | 0.9 | 1 |
|  | ICU: IMV+/-ECMO | 0.24 | 0.4 | 1 | 1 |

The rationale behind the judgements used in the population of the mortality rate assumptions for each care category and scenario are given in Table 2. They were made by Dr Chris Martin a former clinician and the principal architect of this sub-model in conjunction with Dr Rahul Sarkar, Respiratory Physician and Critical Care Consultant at the Medway Hospital Foundation NHS Trust.

**Table 2.** Expert judgements made on mortality rates by care category in the excess mortality sub-model

|  | **Assumption within capacity** | **Assumption with no ICU beds** | **Assumption with no ICU or ward beds** |
| --- | --- | --- | --- |
| **General ward care** | Very low mortality as they are un-escalated, so will only occur with non-respiratory failure deaths or sudden deaths. | Very low mortality as they are un-escalated, so will only occur with non-respiratory failure deaths or sudden deaths. | The risk remains low but will be somewhat greater than the ward-based care. |
| **Ward care O<sub>2</sub>>35%** | This is a group of people, who would be escalated to ICU if they deteriorated, so will only include sudden deaths. | Those who deteriorate may get CPAP on the ward, so use the CPAP mortality from ICU. | Assume substantial mortality as these are very high flow rates indicating severe disease and risk to life. Assume that there will be no O<sub>2</sub> outside of hospital. Set at 50% after discussion. |
| **Ward care O<sub>2</sub>>35% (Ceiling)** | From AW et al. Assumption that this cohort 2 group will be frail with a CFS of 6-9. From figure 1, the mortality is about 40% at 28 days.(11) | Assume unaffected by ICU absence. | This is a group of frail people who will have no medical support despite needing unusually high O<sub>2</sub> flows to remain stable. Assume there will be no O<sub>2</sub> outside of hospital. |
| **Supportive/HFO** | Assume the same as for CPAP. | Assume the same as for CPAP. | An assumption that no O<sub>2</sub> available outside hospital. Mortality likely to be high without it in this group identified at having a risk to life by their admission to ICU. |
| **NIV/CPAP** | In Sivaloganathan et al, there were 31 patients who received NIV only on ICU, none of whom died suggesting the rate is below 3-4%. Deaths are only likely to occur if from non-respiratory causes such as sepsis or non-respiratory organ failure, otherwise they would have been escalated to IMV/ECMO.(10) | Assume that CPAP can continue on the ward but double the mortality for reduces surrounding support etc. | Assume the great majority would die without CPAP, otherwise they would have been be left on O<sub>2</sub>. Also assume that there will be no O<sub>2</sub> outside hospital. |
| **NIV/CPAP (Ceiling)** | Observation from Sivaloganathan et al.(10) | Assume that CPAP can continue on the ward, but with an increase due to a reduction in other supportive care. | Assumption that in the absence of any hospital bed, then there would be no NIV and no chance of survival. |
| **IMV+/-ECMO** | ICNARC Report 30/10/20 - mortality in those ventilated within 24hrs after admission is 40% since 1st Sep. Assumption that the mortality rate is similar for those mechanically ventilated after the 1st 24hrs.(12) | Reasonably safe assumption. IMV/ECMO are dangerous but potentially lifesaving procedures only carried out if considered life-saving in this circumstance. | Reasonably safe assumption. IMV/ECMO are dangerous but potentially lifesaving procedures only carried out if considered life-saving in this circumstance. |

The expected numbers of deaths in each category calculated using the assumed mortality rate and weighted by the proportions in the categories using the formulae below.

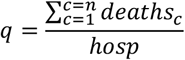

Where:

*q* = probability of dying in this episode of COVID-19

*deaths*_*c*_ = the number of deaths in the care category ‘c’.

*hosp* = the number of people hospitalised with COVID-19.

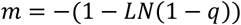

Where:

*m* = hazard rate for death corresponding to the probability of dying in the scenario, and

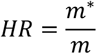

where *HR* = hazard ratio applied to *m* to find the mortality rate in scenario ‘*’.

We estimated a hazard ratio of 1.99 for mortality in those needing ICU care when no ICU bed was available but there was a ward bed and 9.02 when there is neither an ICU nor ward bed available. For those needing ward care only, the hazard ratio is 3.69 when no hospital bed is available. Sensitivity testing indicated that results were not especially sensitive to small changes in these hazard ratios (+/-20%).

### Ward Mortality – Treated

In this step, we estimated the in-capacity death rate by age in 5-year age bands interpolating from data from the cumulative COVID-19 daily deaths report on the NHS England website and using the age distributions for admissions calculated in the modelling step “Estimating Admissions by Age”.(13) Data was harvested from the 6^th^ November 2020 dataset as this precedes the peak of the second wave when pressure may already have been building on internal hospital resources. After estimating the mid-points of the age-bands, an exponential model is fitted to the death rates for the three age bands from age 40 upwards. This fitted model is then used to interpolate the death rates for the 5-year age bands required.

### ICU Mortality – Treated

In this step, we calculated the in-capacity death rates by 5-year age bands in ICU. Data was taken from the 6^th^ November 2020 ICNARC report on COVID-19 in intensive care.(14) The death rate for each age band was calculated from the sum of the product of the number of admissions for that age band, the 28-day in-hospital mortality rate and the total number of admissions for that age band. An exponential model was then fitted to the data to which age bands had been applied in order to allow interpolation and re-categorising by 5-year age-bands.

### Mortality Rates

In this step we took the mortality rates by age calculated in the previous two modelling steps and the hazard ratios from the ‘Excess Mortality Model’ step to calculate the mortality rates when no ICU beds and no ward beds are available respectively. The mortality rate under the scenario ‘*’ is calculated using the equation:

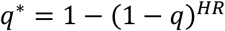

Where:

*q* = the death rate in the in-capacity scenario.

*HR* = the hazard ratio in scenario ‘*’.

*q*\* = the death rate in the scenario ‘*’.

### Deaths - In capacity

In this step, we calculated the number of deaths each week from the corresponding number of admissions generated by the ‘Ward Demand’ and ‘ICU Demand’ steps and the in-capacity mortality rates from the previous ‘Mortality Rates’ step. Observed data is used up to the 9^th^ December 2020, with ICU deaths calculated as 15% of the total number of deaths (as there are significant delays in reporting the deaths in the ICNARC data).

### Deaths - Outside capacity

In this step, we calculated the numbers of deaths arising each week as a direct result of failing to get an ICU or ward bed. Observed data is used up to the 9^th^ December 2020. A series of seven calculations were used:

1. The number of deaths in ICU within capacity as the dot product of the vector of ICU demand within ICU capacity by age from step ‘ICU Demand’ and the vector of mortality rates from step ‘Mortality Rates’.
2. The number of deaths in patients requiring ICU care where only ward care was available, as the dot product of the vector of ICU demand outside ICU by age from step ‘ICU Demand’ and the vector of mortality rates from step ‘Mortality Rates’.
3. The number of deaths in patients requiring ICU care where neither ward nor ICU care was available as the dot product of the vector of ICU demand outside both ICU and ward by age from step ‘ICU Demand’ and the vector of mortality rates from step ‘Mortality Rates’.
4. The number of deaths in patients requiring ward care only as the dot product of the vector of ward demand within ward capacity by age from step ‘Ward Demand’ and the vector of mortality rates from step ‘Mortality Rates’.
5. The number of deaths in patients requiring ward care where ward care was not available as the dot product of the vector of ward demand outside ward capacity by age from step ‘Ward Demand’ and the vector of mortality rates from step ‘Mortality Rates’.
6. The total number of deaths each week under the current scenario including both ICU and ward deaths as the sum of all results in steps 1-5 above.
7. The total number of deaths in each calendar week in both the ICU and ward care groups calculated in the step ‘Deaths - In capacity’ was taken and subtracted from the capacity constrained total in step 6 above to provide the number of deaths arising as a direct result of the lack of an ICU or ward bed.

### Scenarios

We applied three scenarios for admission demand from the week beginning 16^th^ December 2020 through to the week beginning 28^th^ April 2021 (Figure 2). In this model, the three scenarios are referred to as “Pessimistic”, “Middling” and “Optimistic” and were arbitrary but based on the trajectory of the increase in the multiplier of admissions from one week to the next. The trajectory was calculated as a linear regression of the last three weeks of the known data taken from the GOV.UK Coronavirus dashboard.(7) They are illustrative scenarios and not forecasts.

**Figure 2.**
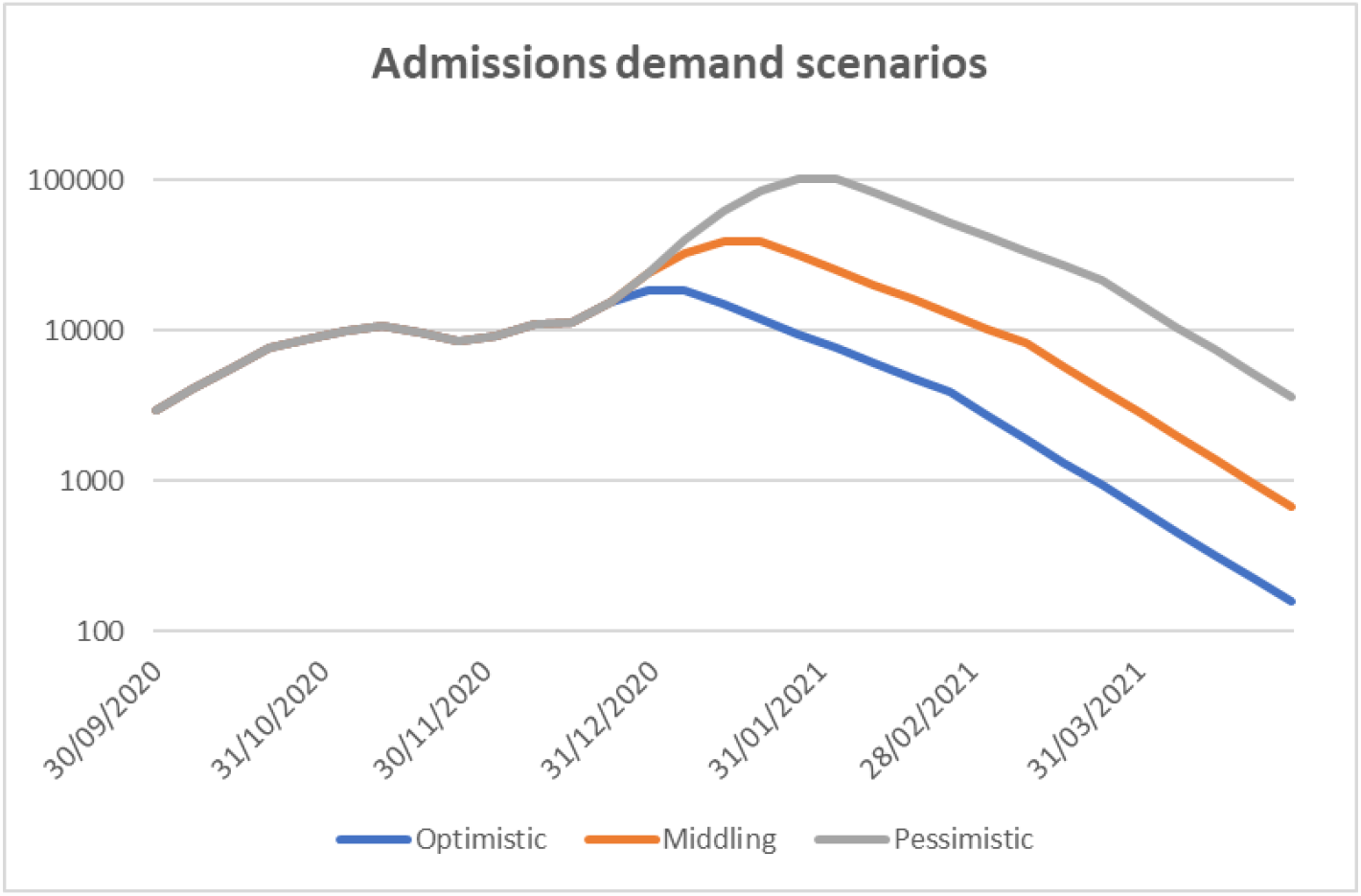
Graph showing three scenarios of projected admission demand from the 9th December 2020.

In the Middling scenario, the multiplier continued to increase following the rate over the previous 3 weeks up to three weeks post the introduction of more vigorous countermeasures on 19^th^ December 2020. This is to be expected as the incubation period is nearly a week, so the enhanced countermeasures would usually take at least a week to result in any observed change. The multiplier falls below 1.0 after 4 more weeks. The emergence of a new variant of SARS-CoV-2 with higher transmission rates that became more prevalent from the beginning of December has driven more rapid growth of infection rates. As the new variant makes up a greater proportion of new cases, the rate of growth will continue to rise unless effectively mitigated. It remains uncertain at the time of writing how it will respond to tighter controls, but there is no reason to assume the mortality rates will differ at present. At the time of writing this new variant has only recently been identified and has not been factored into the analysis. However, initial indications are that this variant could open up a variety of more pessimistic scenarios in the short term.

In the Optimistic scenario, the multiplier begins to decelerate from week beginning 16^th^ December, continues to decelerate after the 16^th^ December and falls below 1.0 after 3 weeks.

In the Pessimistic scenario, the multiplier continued to increase for 4 weeks after week beginning 16^th^ December before decelerating at the same rate as the other two scenarios. This is not intended as a worst-case scenario, but one that is somewhat worse than the ‘Middling’ scenario.

### Sensitivity analysis

We performed an analysis to assess the sensitivity of the model to the choice of capacity and hazard ratio parameter values and scenario. Five input parameters were varied independently in turn and the effect on the model output recorded. The five parameters varied were the ICU and ward capacities and the hazard ratios for mortality in ward patients when there are no ward beds available and ICU patients when there are no ICU beds and ICU patients when there are no ward beds available. The hazard ratios were adjusted using the equation.

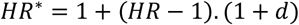

Where:

*HR* = the within-capacity hazard ratio.

*HR*^*^ = the outside-capacity hazard ratio.

*d* = the adjustment to be applied as a proportion.

This model has been populated with data specific to England, it could be applied in other geographies and we have added ways of estimating some data like the age distribution of admissions or the ICU admission rates by age. Where this data is directly available it can be substituted.

### Assumptions

There are several general underlying assumptions to the model.

- There is no change in the expected mortality as demand grows until the bed capacity is breached.
- Only bed capacity affects excess mortality rates and not other resource constraints such as staffing, equipment, ambulance availability or other finite resources.
- Health care policy and conscious or unconscious clinician behaviour that may affect the compartments of care through which the patients flow does not change as the pandemic waxes and wanes.
- Surge capacity will be applied before the use of field hospitals or the commandeering of private facilities.
- All the surge capacity, field hospital and private facility beds will be utilised for COVID-19 rather than any other demand.
- The age distribution of admissions remains constant over time.
- There is perfect redistribution of bed capacity and resources across the country immediately according to demand.
- Weekly bed availability remains constant.
- Capacity estimates are fixed throughout the model when, in fact, new resources may be recruited over time, or capacity may fall in response to disruption of staffing and supplies.
- The mortality rates, including by age, are the same for the old COVID strain and novel mutations.

## Results

### Sensitivity analyses

*The results of the sensitivity analysis are shown in* Table 4 and **Error! Reference source not found**.. None of the Optimistic scenario adjustments resulted in any excess deaths, and in the Middling scenario adjustments, the excess deaths attributable to lack of capacity alone ranged from 0 to 597 with a 20% reduction in the expected number of ward beds. In the Pessimistic scenario the number of excess deaths attributable directly to lack of capacity range from 49,219 with a 20% increase in the number of ward beds, and 103,845 with a 20% reduction in the expected ward bed availability.

**Table 3.** Numbers of deaths with the unadjusted middling scenario from 9th of December.

| Week | Total Deaths (with ICU and Ward capacity limits) | Deaths if there are no ICU and Ward capacity limits. | Excess deaths specifically due to ICU and Ward capacity limits |
| --- | --- | --- | --- |
| 30/09/2020 | 298 | 298 | 0 |
| 07/10/2020 | 444 | 444 | 0 |
| 14/10/2020 | 731 | 731 | 0 |
| 21/10/2020 | 1,074 | 1,074 | 0 |
| 28/10/2020 | 1,457 | 1,457 | 0 |
| 04/11/2020 | 1,784 | 1,784 | 0 |
| 11/11/2020 | 1,897 | 1,897 | 0 |
| 18/11/2020 | 2,103 | 2,103 | 0 |
| 25/11/2020 | 2,031 | 2,031 | 0 |
| 02/12/2020 | 1,889 | 1,889 | 0 |
| 09/12/2020 | 1,960 | 1,960 | 0 |
| 16/12/2020 | 2,330 | 2,330 | 0 |
| 23/12/2020 | 3,214 | 3,214 | 0 |
| 30/12/2020 | 4,945 | 4,945 | 0 |
| 06/01/2021 | 8,399 | 8,399 | 0 |
| 13/01/2021 | 10,156 | 10,078 | 77 |
| 20/01/2021 | 10,156 | 10,078 | 77 |
| 27/01/2021 | 8,063 | 8,063 | 0 |
| 03/02/2021 | 6,450 | 6,450 | 0 |
| 10/02/2021 | 5,160 | 5,160 | 0 |
| 17/02/2021 | 4,128 | 4,128 | 0 |
| 24/02/2021 | 3,302 | 3,302 | 0 |
| 03/03/2021 | 2,642 | 2,642 | 0 |
| 10/03/2021 | 2,114 | 2,114 | 0 |
| 17/03/2021 | 1,479 | 1,479 | 0 |
| 24/03/2021 | 1,036 | 1,036 | 0 |
| 31/03/2021 | 725 | 725 | 0 |
| 07/04/2021 | 507 | 507 | 0 |
| 14/04/2021 | 355 | 355 | 0 |
| 21/04/2021 | 249 | 249 | 0 |
| 28/04/2021 | 174 | 174 | 0 |
| **Total** | **91,251** | **91,096** | **154** |

**Table 4.** Results of the sensitivity analysis.

| Scenario | Ward availability | ICU availability | No ward HR | No ICU HR | No ICU or Ward HR | Excess deaths due to capacity breach | Total deaths | % increase in deaths |
| --- | --- | --- | --- | --- | --- | --- | --- | --- |
| Middling | -20% | 0% | 0% | 0% | 0% | 154 | 91,251 | 0.2% |
| **Middling** | **0%** | **0%** | **0%** | **0%** | **0%** | **154** | **91,251** | **0.2%** |
| Middling | 20% | 0% | 0% | 0% | 0% | 154 | 91,251 | 0.2% |
| Middling | 0% | -20% | 0% | 0% | 0% | 597 | 91,693 | 0.7% |
| Middling | 0% | 20% | 0% | 0% | 0% | 0 | 91,096 | 0.0% |
| Middling | 0% | 0% | -20% | 0% | 0% | 154 | 91,251 | 0.2% |
| Middling | 0% | 0% | 20% | 0% | 0% | 154 | 91,251 | 0.2% |
| Middling | 0% | 0% | 0% | -20% | 0% | 130 | 91,226 | 0.1% |
| Middling | 0% | 0% | 0% | 20% | 0% | 176 | 91,272 | 0.2% |
| Middling | 0% | 0% | 0% | 0% | -20% | 154 | 91,251 | 0.2% |
| Middling | 0% | 0% | 0% | 0% | 20% | 154 | 91,251 | 0.2% |
| **Optimistic** | **0%** | **0%** | **0%** | **0%** | **0%** | **0** | **42,925** | **0.0%** |
| Pessimistic | -20% | 0% | 0% | 0% | 0% | 103,845 | 357,600 | 40.9% |
| **Pessimistic** | **0%** | **0%** | **0%** | **0%** | **0%** | **73,783** | **327,538** | **29.1%** |
| Pessimistic | 20% | 0% | 0% | 0% | 0% | 49,219 | 302,975 | 19.4% |
| Pessimistic | 0% | -20% | 0% | 0% | 0% | 76,454 | 330,210 | 30.1% |
| Pessimistic | 0% | 20% | 0% | 0% | 0% | 71,325 | 325,081 | 28.1% |
| Pessimistic | 0% | 0% | -20% | 0% | 0% | 65,912 | 319,667 | 26.0% |
| Pessimistic | 0% | 0% | 20% | 0% | 0% | 80,662 | 334,418 | 31.8% |
| Pessimistic | 0% | 0% | 0% | -20% | 0% | 73,544 | 327,299 | 29.0% |
| Pessimistic | 0% | 0% | 0% | 20% | 0% | 73,998 | 327,753 | 29.2% |
| Pessimistic | 0% | 0% | 0% | 0% | -20% | 73,015 | 326,770 | 28.8% |
| Pessimistic | 0% | 0% | 0% | 0% | 20% | 74,269 | 328,024 | 29.3% |

The results can be explained by considering how capacity evolves in each of the scenarios. In the Middling scenario, whilst ICU capacity may be approached and even possibly breached, there remains sufficient ward capacity to take lives who need either ward or ICU support, keeping excess deaths relatively low. However, the Pessimistic scenario sees ward capacity breached, and in many scenarios for a period of several weeks, resulting in much higher mortality in those lives who require care but do not receive it. ICU capacity is much lower than ward capacity and only a small proportion of all hospitalized patients need ICU care so the number of deaths from breaches of ward capacity are proportionally larger than breaches for ICU care. ICU care is assumed to reach capacity before ward care, so with marginal breaches of capacity, ICU breaches are the source of the excess deaths, but with large breaches of capacity, the majority will arise from breaching of ward capacity.

The number of excess deaths is most sensitive to ward bed availability in the pessimistic scenario with a difference of 54,626 between the 20% increase and 20% decrease in the bed availability compared to 5,129 with the same variation in ICU bed availability. However, in the Middling scenario, the excess deaths are most sensitive to ICU bed capacity with a difference of 597 with +/-20% variation in the ICU bed capacity estimate, with no difference arising from the ward bed capacity estimate.

These results are shown graphically in Figure 3. (Please note the different y-axis scales). Excess deaths are clearly more sensitive to the availability of ICU or ward beds than to the adjustments in the hazard ratios used here. The greatest loss of life occurs with a 20% reduction in the estimate of ward availability in the pessimistic scenario with 103,845 (40.9% increase) excess deaths.

**Figure 3.**
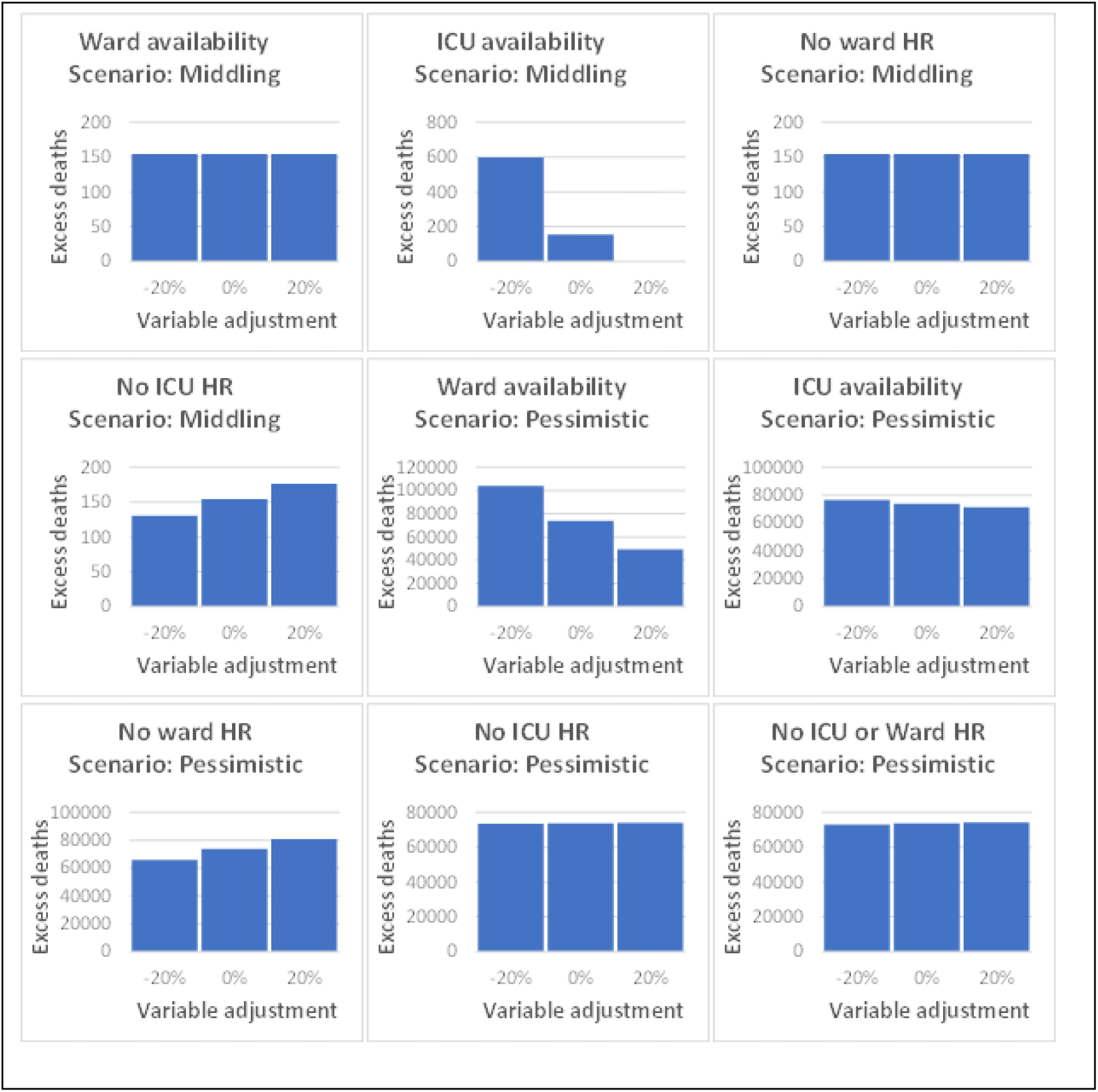
Results of the sensitivity analysis.

## Discussion

In none of the Optimistic scenarios are there any deaths that occur as a direct result of lack of capacity. Several of the ‘Middling’ scenario applications under sensitivity testing resulted in excess deaths directly attributable to a lack of capacity. Most excess deaths arose when we modelled a 20% reduction comparted to best estimate ICU capacity. This led to 597 deaths (0.7% increase).

All the ‘Pessimistic’ scenario applications under sensitivity analysis had excess deaths. These ranged from 49,219 (19.4% increase) when we modelled a 20% increase in ward bed availability over the best-estimate, to 103,845 (40.9% increase) when we modelled a 20% shortfall in ward bed availability below the best-estimate.

The Middling scenario entails a three-week projection of the rate of increase in the multiplier of cases observed between 25^th^ November 2020 and 9^th^. This would be consistent with the lag that occurs because of the approximate one-week incubation period and testing lag from the 19^th^ December 2020 when more restrictive social distancing measures were introduced in the form of Tier 4. A steady decline is then applied so that the multiplier falls below ‘1’ after four weeks as may be expected after their introduction. In the Pessimistic scenario, the rise in the multiplier continues until the first week of January 2021 as might occur if there is poor compliance with social distancing over the Christmas holiday.

The new variant of SARS-CoV-2 that emerged in the UK (VOC 202012/01) appears to be about 56% (95% CI 50%-74%) more transmissible than the existing variants though there is no evidence that the virulence is any different.(15) Alternative explanations for its rise in prevalence and the increased rate of transmission observed since its appearance at the beginning of October were investigated including “immune escape” where individuals previously infected return to susceptibility as a result of mutation of key antigens, increased susceptibility amongst children, and a shorter generation time. None of these alternative explanations fitted the data as well as increased infectiousness. In addition, a second variant has emerged from South Africa (501Y.V2) which also appears to have greater transmissibility and an increased viral load.(16) These unsettling developments increases uncertainty in the future trajectory of hospital demand and open up a variety of significantly more pessimistic scenarios in the short term that have not been possible to explore here.

### Limitations

Here we list and discuss the limitations of the model:

- The estimate of bed availability is determined using the mean length of stay on the ward or in ICU. The distribution of occupants by length of stay will change over time which may result in a slow consumption of capacity that is not captured in this model.
- Mortality rates are affected by constraints other than just bed availability including staffing and equipment.
- The model will not capture the transition between low mortality with full capacity and the high mortality from lack of a bed that arises from stressing of the system before capacity is absent.
- Capacity estimates are fixed throughout the model when, in fact, new resources may be recruited over time, or capacity may fall in response to disruption of staffing and supplies.
- Age specific case fatality rates are assumed to be static, but in fact may change, either due to changes in virulence or improvement in care.

Intensive care resources are constrained not only by bed availability but also by equipment and staffing. A combination of all three is required for optimal care in ICU. In reality there isn’t a simple binary state of presence or absence of these factors and skills. A degradation of equipment maintenance and distribution as well as reductions in the effectiveness of staff, either because staff to patient ratios fall as demand rises, or because of staff sickness due to COVID-19 or simply the physical and emotional fatigue as the pandemic continues is also seen. The modelling by McCabe et al suggests that ICU capacity is first constrained by bed availability, though lack of nurses and junior doctors is close behind.(2) They included sickness absence rates taken from surveys of union members suggesting 15% of doctors were off sick in the first wave and may under-estimate the impact of sickness on staffing overall at peak times in the pandemic.(17) As the pandemic progresses, higher than anticipated absence due to sickness in these groups could result in lack of limiting capacity before the lack of beds. Reorganisation within hospitals may mitigate this by training other staff to support ICU work and thereby increasing ICU staff to bed ratios.(18) One hospital managed to meet demand in the first wave, but reduced ICU nurse to bed ratios from the normal pre-pandemic of 1:1 to 1:4.(4) Current guidance on nursing staff ratios during the COVID-19 pandemic advocates a ratio of 1:2.(18)

Decision making may change in the face of increasing demand either with formal revisions of treatment thresholds as resources become increasingly scarce or with the introduction of triaging. On a less formal level, the heuristics used by clinicians in their everyday management of patients may vary as competing pressures rise. For example, at times of abundant capacity, the thresholds for transferring patients into ICU for a trial to see if a patient with poor chance of survival recovers with a short stay, may be lower than when there is severe limitation on capacity.

An important but unrealistic assumption in the model is that there is perfect distribution of resources with respect to demand across the country and that no patient is refused care whilst there remains a bed anywhere in England. ICUs are organized into networks which facilitate the transfer of patients from one hospital to another when ICU beds in a hospital run out, but even a delay of a few hours in transferring a patient to an ICU bed can influence outcome. Furthermore, even in the case of bed availability in a different hospital, it cannot be assumed that the patient in the referring and at-capacity hospital may be fit enough to able to be transferred safely. All the ICUs in a network are likely to have correlated demand and may all run out of space at the same time. In these circumstances, transfers would need to be arranged between networks or regions and this would entail yet further delay and challenges with consequent impact on outcomes.

It is notable that the sensitivity of the excess deaths to ICU capacity is much greater than it is to ward bed capacity at marginal breaches of capacity, but that large scale breaches of capacity are more sensitive to ward bed capacity. There are far more ward beds than ICU beds, and there is an assumption that ICU capacity would be exhausted long before ward bed capacity.

## Conclusions

Here we describe a demand and capacity model for general hospital and intensive care beds in the context of the COVID-19 pandemic in England. The model allows a user to understand the excess mortality impact arising as a direct consequence of capacity being breached under various scenarios or forecasts of hospital admissions. The scenarios described in this paper are illustrative and are not forecasts.

We estimated the number of excess deaths up to the end of April 2021 that would arise from lack of capacity under different demand assumptions from the 16^th^ December 2020. No excess deaths from excess capacity would be expected under the ‘Optimistic’ assumptions of demand but would reach between 49,219 and 103,845 under the ‘Pessimistic’ scenario. Without the new variant, exceeding capacity for hospital and ICU beds was not the most likely outcome, but given the new variant it now appears more plausible and if so would result in a substantial increase in the number of deaths from COVID-19.

## Supporting information

Excel spreadsheet of the model.

## Data Availability

The model is available on GitHub here:

https://github.com/Crystallize/COVID19_ExceedingCapacityModel

## List of abbreviations

HFO: high flow oxygen
NIV: non-invasive ventilation
CPAP: continuous positive airway pressure
IMV: invasive mechanical ventilation
ECMO: extracorporeal membrane oxygenation

## Declarations

### Ethics approval and consent to participate

Not appliable

### Consent for publication

Not applicable

### Availability of data and materials

*Will probably require a statement (website has various template statements we can use depending on type of dataset/materials)*

### Competing interests

The authors declare that they have no competing interests.

### Funding

No funding source.

## Authors’ contributions

Stuart McDonald initiated and co-ordinated the project and was one of the principal architects of the general model and contributed to the final paper.

Chris Martin was the principal architect of the excess mortality sub-model, participated in the development of the general model and pulled together the initial draft of the paper.

Michiel Luteijn participated in the development of the general model and undertook ancillary modelling that supported development and contributed to the final paper.

Steve Bale participated in the development of the general model and contributed to the final paper.

Rahul Sarkar contributed to the expert judgements in the modelling, advised on the organizational context within which the model sits, and contributed to the final paper

## Acknowledgements

**Various**

Colin Dutkiewicz, (Global Head of Life Reinsurance Solutions, Aon)

Josephine Robertson, (Health & Care Actuary, Optum)

Dan Ryan, (Chief Science Officer at COIOS Research)

Scott Reid, (Global Protection Pricing & Product Development Actuary, Zurich)

James Robinson, (UK & Ireland Business Strategy and Analytics, Aon Benfield)

**Nottingham Universities Hospital Trust**

Joe West, (Professor of Epidemiology; Honorary Consultant Gastroenterologist, Faculty of Medicine & Health Sciences Nottingham University)

Richard Hubbard, (GSK/British Lung Foundation Professor of Respiratory Epidemiology, Faculty of Medicine & Health Sciences)

Tim Card, (Clinical Associate Professor, Nottingham Biomedical Research Centre)

Colin Crooks, (Clinical Associate Professor, Nottingham Biomedical Research Centre)

Nina Lewis, (Consultant Gastroenterologist, Nottingham University NHS Trust)

Andrew Fogarty (Clinical Associate Professor & Reader in Clinical Epidemiology, Faculty of Medicine & Health Sciences)

Andrew Marshall, (Deputy Medical Director at Nottingham University Hospitals)

De Beer Thearina, (Consultant in Anaesthetics and ICM at Nottingham University Hospitals NHS Trust)

Dominic Shaw, (Professor and Honorary Consultant, Faculty of Medicine & Health Sciences)

Emma O’Dowd, (Honorary Consultant Assistant Professor, University of Nottingham)

Andrew Baraclough, (Assistant Director of Insight, Nottingham University Hospitals NHS Trust)

**Crystallise**

Shannon Connolly, Analysts at Crystallise Ltd.

Dr Will Letton, Consultant Analyst at Crystallise

Dr Louisa Rutherford, Senior Clinical Researcher at Crystallise Ltd.

## Notes

### Competing Interest Statement

The authors have declared no competing interest.

### Funding Statement

There was no funding provided.

